# Views of the Swiss public towards gene editing

**DOI:** 10.1101/2025.09.28.25336849

**Authors:** KE Ormond, S Kijewski, N Wyler, A Ferretti, E Petrou, C Reichmuth, E Vayena

## Abstract

There is little country-specific data about how the general public views gene editing therapies. In Autumn 2023 we randomly surveyed the Swiss public, using the Federal Register and stratifying by language region (German, French, Italian), gender and age. We present a representative sample of 3855 responses, including >4000 open-ended comments. When presented with 7 therapeutic options for somatic gene editing, 7% disagreed with all therapeutic options, and 35% supported them all. Most agreed with using somatic gene editing to cure life-threatening (76%) and debilitating diseases (70%); support declined as severity decreased or with later onset. Few supported somatic gene editing to enhance physical (6%) or cognitive (9%) abilities. In all scenarios, people were less likely to agree with gene editing of embryos. Notably, all therapeutic gene editing attitudes clustered, regardless of somatic vs. germline differentiations. Factor analysis also demonstrated two clusters for “support” and “caution” towards gene editing, and multivariate analysis demonstrated relationships with age, gender, religion and knowledge. When asked what influenced their views, the most endorsed reasons for feeling positively were ‘views towards what it means to have a good life’ (59.8%) and ‘views about illness and suffering’ (58.5%). Most selected “neutral” to describe religion’s influence (68.9%) despite findings that those who endorsed high religiosity were less supportive and more cautious towards gene editing. Conclusions: Uncovering systematic differences in the attitudes towards specific therapies and the values shaping them underscores the importance of including peoples’ voices in policy decisions in a country-specific manner.

## Introduction

Discussions of public views towards gene therapies and gene editing have been published since the early 1990s, but their frequency has increased in the past decade as CRISPR-Cas9 and newer therapies have made the option more technologically feasible in the near term. There is a range of papers that provide insight to the public’s views, many reviewed by Delhove et al (2020)^1^, but they are challenged by covering only germline gene editing,^2–5^ using different methodologies (survey versus public engagement events, for example), or phrasing questions in different (non-validated) manners. Additionally, several were published in 2016-2017,^6,7^ when the technology was still very hypothetical.

Nevertheless, some data exists describing views of the general public towards gene editing from the US, Canada, UK, EU, Netherlands, South Africa, Japan, China and Australia. Papers assessing the general public generally show greater support for somatic gene editing (SGE) than for germline gene editing (GGE)^1,7–9^. Only two large public studies show non-significant differences between these views^6,10^. Studies also generally agree that support increases with disease severity^2,4,6,8,11–13^ and decreases when considering enhancement (for example of physical or cognitive features)^4–7^. Importantly, some significant part of the population often disagrees with any use - for example in Japan up to 30% of the general public were not supportive^8,14^.

Finally, factors that seem to influence public views have also been examined, including as part of the Delhove review. Most studies that examined religiosity found that those who self-describe as more religious have lower support for gene editing^1,5,6,10,12,13^, particularly for GGE^3^. When age played a role, younger individuals were more supportive^1,5^. Education level was associated with higher support for gene editing^1,12,13,15^. When gender differences were found, men were more supportive ^1,5,6,8,10,16^. Personal or family history of genetic disease showed mixed findings: some studies suggested this did not influence views towards gene editing^1,13^, but studies of patient and family stakeholders identified specifically around genetic disease groups showed higher interest ^4,17–21^.

Several authors comment that country-specific data will be useful to gather given the important differences noted thus far. Country-specific data can guide policy creation in a way that recognizes important values and cultural differences that may not otherwise appear. In Switzerland, a small country with high literacy rates^22^, we recently collected data on how scientific and medical professionals in Switzerland view gene editing^23^, but what is not known is how the Swiss public thinks about the potential for gene editing. As such, we constructed the current study to assess the attitudes of the Swiss public towards gene editing, including how these attitudes compare to the already assessed public views.

## Materials and Methods

We conducted an online cross-sectional survey using Qualtrics to learn about the opinions, concerns and expectations of Swiss residents towards gene editing.

### Ethics approval statement

The survey was approved by the ETH Zurich Ethics Commission (EK 2022-N-84). Participants reviewed the informed consent materials before completing the survey, which included the statement “By participating in the survey, you are giving your informed consent to participate in this research study.”

### Subjects

Potential participants were 18-72 years old and lived in Switzerland. In order to best represent the general public of Switzerland, the Swiss Federal Statistical Office (FSO) provided a stratified random sample across sex, age (4 groups, between 18 and 64+), and three main language regions (German, French, Italian) in Switzerland. Since a majority (62%) of the Swiss population speaks primarily German, and only 23% and 8% speak French and Italian, respectively, the minority languages were oversampled ( 2:1:1 for German, French, Italian) in order to maximize responses from these populations. Anticipating an approximately 30% response rate, the target population included 14,825 individuals (6977 German speakers, 4013 French speakers, and 3835 Italian speakers).

### Questionnaire development

The survey (Supplemental Methods, in English) was based on an unpublished survey that was used by the Australian Citizen Jury research team (personal communication, S Niemeyer). Several questions were added after interviews with Swiss experts to obtain their views on gene editing and expectations for areas where the Swiss public might differ uniquely^23^. The survey was divided into four parts. In the first part questions were asked about the participants’ background such as canton of residency, gender, age, education level, religious beliefs and political attitude. In the second part two questions about the awareness and pre-knowledge about gene editing were asked. Then, a short explanation about gene editing in simple language was given to all participants. In the third part, views on human gene editing (18 questions) and its potential usages (15 questions) were investigated. We used a Likert Scale either ranging from 1-5 or 1-7, where 1 = strongly disagree and 5 or 7 = strongly agree. The fourth part contained questions about the attitudes towards the regulation of human gene editing. The operationalization of these variables is described in the Supplemental Methods (Table S1).

The survey was created in English, and then translated to German, French and Italian using DeepL (an artificial intelligence translator) and then error checked and edited for fluency by at least two team members for whom the language was their mother tongue. Non-English versions of the survey are available on request.

### Recruitment

Recruitment mailings were sent between September 7 and October 5 2023. Potential participants were mailed a letter in their preferred language of correspondence that briefly described the study and gave instructions to access the survey, either online or to request a paper version. A secured individualized code that links the data to the sociodemographic data provided by FSO was given in order to be able to anonymously track the individual responses. Two reminder notifications were sent to non-responders at two and four weeks. The data was stored safely in an encrypted form at ETH.

The survey was administered over Qualtrics (Provo, UT; version accessed between June 2023 and June 2024) and participants could toggle between four different languages (English, German, French, Italian) while completing the survey if desired. Forty paper surveys were requested; these were mailed with the secured code and entered by hand by a single researcher. The final sample contained 3855 respondents who completed attitudinal questions beyond demographics.

### Statistical analysis

We analyzed the data using the software Stata (version Stata/SE 17.0 for Mac, StataCorp, College Station). To reduce sampling error and account for non-response bias, we applied post-stratification survey weights in the data analysis that were calculated based on gender, age and language region. To examine variation in individual views on gene editing, the data was examined using ordinary least squares-regression analysis. For this analysis, we created factor variables estimated based on maximum-likelihood factor analysis of a polychoric correlation matrix, given the ordinal nature of the variables. Our analysis included age, gender, level of education, religiosity, whether the respondent or a family member has an inherited or genetic condition, experience in medical or related fields, political participation, level of knowledge on gene editing, language and region. Further, we included data provided on household size, marital status and nationality acquired from the FSO.

### Qualitative analysis

The survey contained 5 open-ended questions that resulted in 4095 comments, which were qualitatively analyzed using thematic analysis^24^. In order to get an overview of the mentioned themes, select fluent members of the research team (KO, NW, CR, EP, AF) reviewed comments in the original languages to inductively create a draft code book and definitions. The codebook was then tested using 30 English comments (since all researchers were fluent in English). After codebook revisions, a second training round with 10 English comments was conducted. The final code book was then used to analyze the data in NVivo (version 20). To maximize rigor and inter-rater reliability, 20% of the comments were co-coded by two members. The codes were thematically analyzed to understand the main themes and to assist in interpreting the quantitative study results.

## Results

### Sample characteristics

298 survey invitations were returned due to incorrect addresses and 23 persons responded to refuse survey participation. Of the remaining potential participants, 4171 surveys were started online (response rate = 29%), and 3855 surveys were included in the final sample, with 3429 included in the main regression analysis. Table 1 describes the participants’ demographics in the final sample (n=3855). 82% of participants were Swiss, and the population was approximately 50% women with a mean age 48 ± 16 (SD); 54% were married and 10% self-report they are very religious. All language regions were represented, with an over-representation of participants from the Italian speaking region (Ticino) (33% of responses vs. 4% of population). Twenty-seven percent of the participants had a university degree, and 10% worked in a related field of biology or medicine. Approximately 20% reported an inherited disease in their family.

**Table 1.**
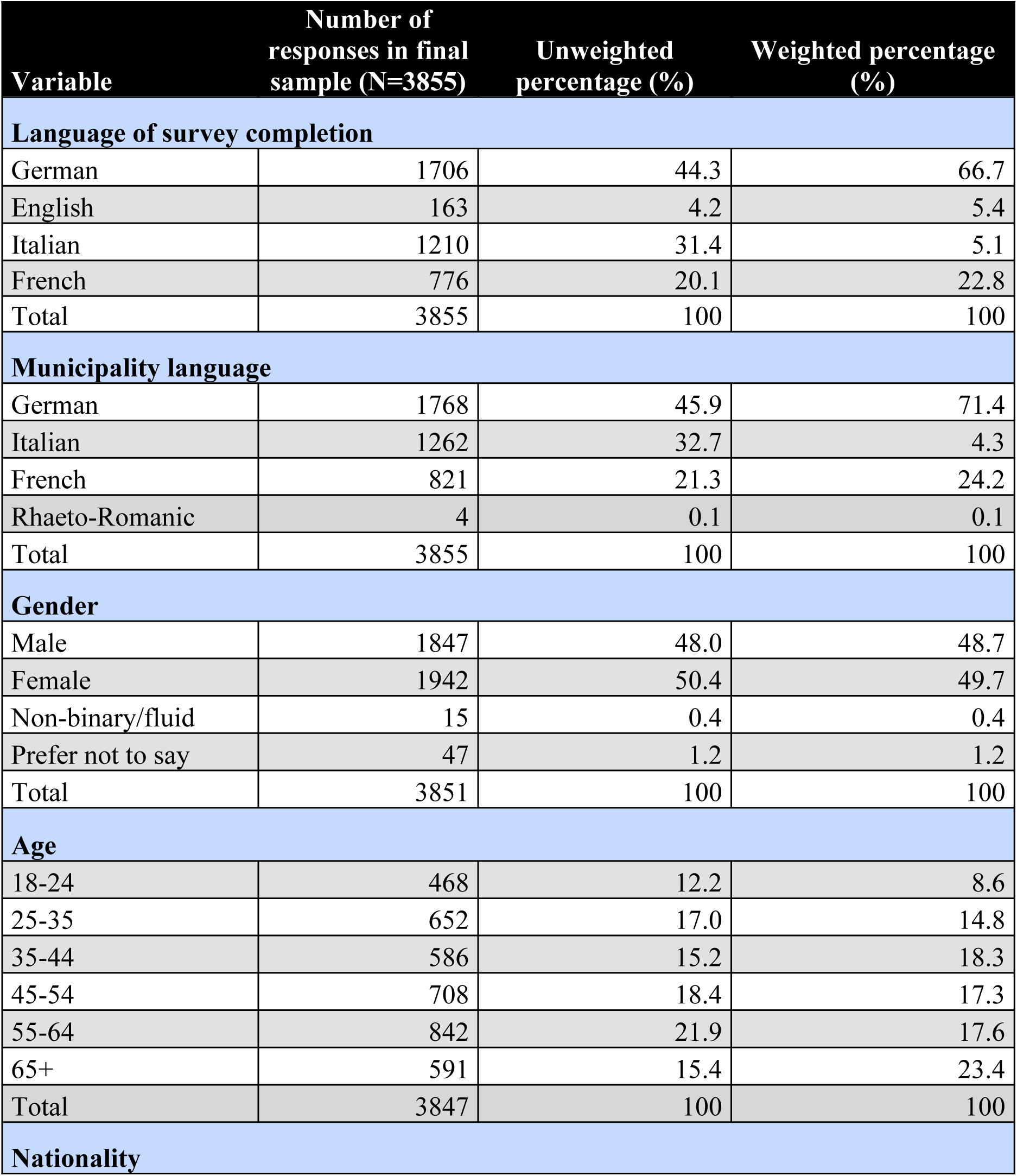
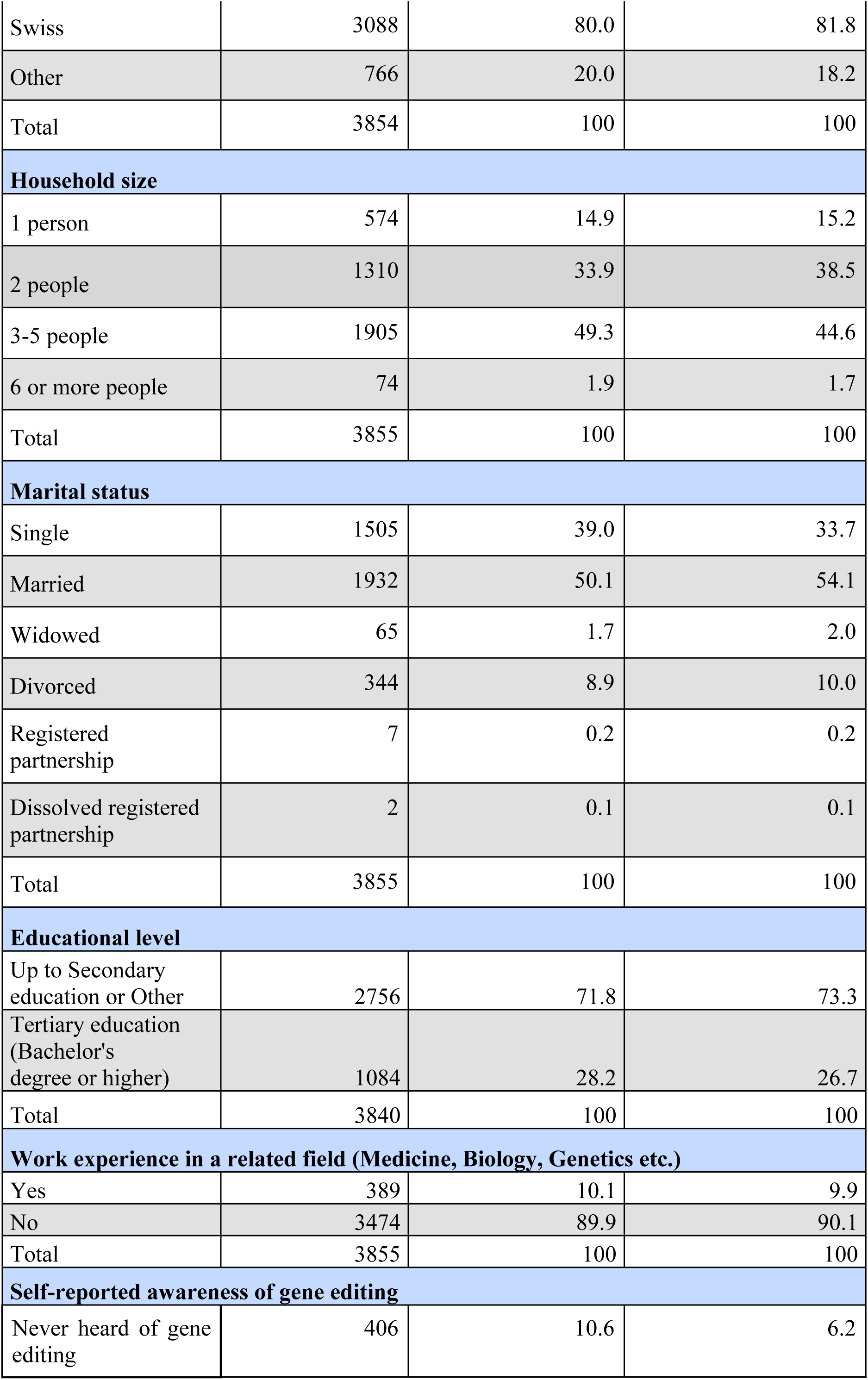
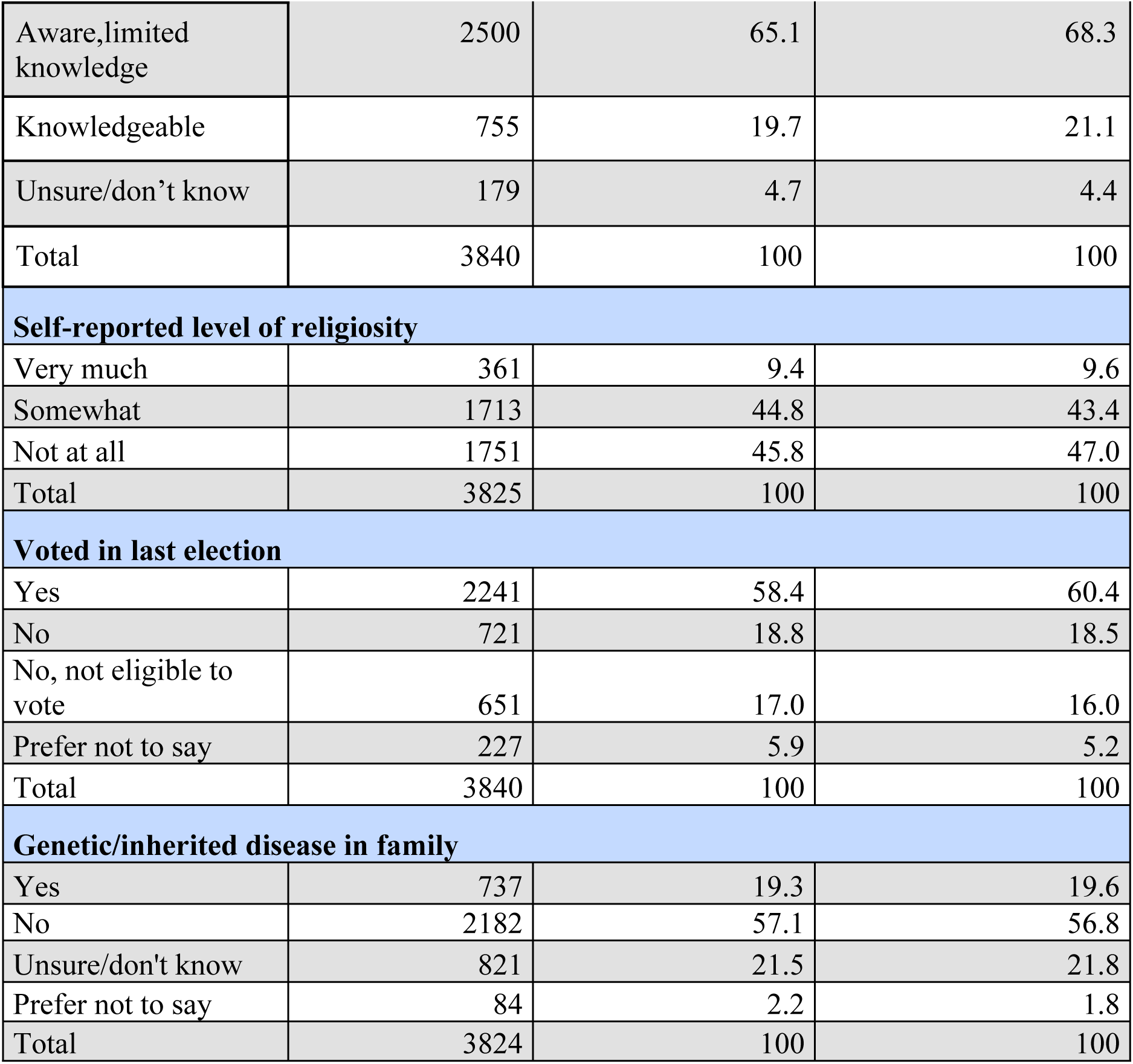
Participant Demographics.

| Variable | Number of responses in final sample (N=3855) | Unweighted percentage (%) | Weighted percentage (%) |
| --- | --- | --- | --- |
| **Language of survey completion** |  |  |  |
| German | 1706 | 44.3 | 66.7 |
| English | 163 | 4.2 | 5.4 |
| Italian | 1210 | 31.4 | 5.1 |
| French | 776 | 20.1 | 22.8 |
| Total | 3855 | 100 | 100 |
| **Municipality language** |  |  |  |
| German | 1768 | 45.9 | 71.4 |
| Italian | 1262 | 32.7 | 4.3 |
| French | 821 | 21.3 | 24.2 |
| Rhaeto-Romanic | 4 | 0.1 | 0.1 |
| Total | 3855 | 100 | 100 |
| **Gender** |  |  |  |
| Male | 1847 | 48.0 | 48.7 |
| Female | 1942 | 50.4 | 49.7 |
| Non-binary/fluid | 15 | 0.4 | 0.4 |
| Prefer not to say | 47 | 1.2 | 1.2 |
| Total | 3851 | 100 | 100 |
| **Age** |  |  |  |
| 18-24 | 468 | 12.2 | 8.6 |
| 25-35 | 652 | 17.0 | 14.8 |
| 35-44 | 586 | 15.2 | 18.3 |
| 45-54 | 708 | 18.4 | 17.3 |
| 55-64 | 842 | 21.9 | 17.6 |
| 65+ | 591 | 15.4 | 23.4 |
| Total | 3847 | 100 | 100 |
| **Nationality** |  |  |  |
| Swiss | 3088 | 80.0 | 81.8 |
| Other | 766 | 20.0 | 18.2 |
| Total | 3854 | 100 | 100 |
| **Household size** |  |  |  |
| 1 person | 574 | 14.9 | 15.2 |
| 2 people | 1310 | 33.9 | 38.5 |
| 3-5 people | 1905 | 49.3 | 44.6 |
| 6 or more people | 74 | 1.9 | 1.7 |
| Total | 3855 | 100 | 100 |
| **Marital status** |  |  |  |
| Single | 1505 | 39.0 | 33.7 |
| Married | 1932 | 50.1 | 54.1 |
| Widowed | 65 | 1.7 | 2.0 |
| Divorced | 344 | 8.9 | 10.0 |
| Registered partnership | 7 | 0.2 | 0.2 |
| Dissolved registered partnership | 2 | 0.1 | 0.1 |
| Total | 3855 | 100 | 100 |
| **Educational level** |  |  |  |
| Up to Secondary education or Other | 2756 | 71.8 | 73.3 |
| Tertiary education (Bachelor's degree or higher) | 1084 | 28.2 | 26.7 |
| Total | 3840 | 100 | 100 |
| **Work experience in a related field (Medicine, Biology, Genetics etc.)** |  |  |  |
| Yes | 389 | 10.1 | 9.9 |
| No | 3474 | 89.9 | 90.1 |
| Total | 3855 | 100 | 100 |
| **Self-reported awareness of gene editing** |  |  |  |
| Never heard of gene editing | 406 | 10.6 | 6.2 |
| Aware,limited knowledge | 2500 | 65.1 | 68.3 |
| Knowledgeable | 755 | 19.7 | 21.1 |
| Unsure/don't know | 179 | 4.7 | 4.4 |
| Total | 3840 | 100 | 100 |
| **Self-reported level of religiosity** |  |  |  |
| Very much | 361 | 9.4 | 9.6 |
| Somewhat | 1713 | 44.8 | 43.4 |
| Not at all | 1751 | 45.8 | 47.0 |
| Total | 3825 | 100 | 100 |
| **Voted in last election** |  |  |  |
| Yes | 2241 | 58.4 | 60.4 |
| No | 721 | 18.8 | 18.5 |
| No, not eligible to vote | 651 | 17.0 | 16.0 |
| Prefer not to say | 227 | 5.9 | 5.2 |
| Total | 3840 | 100 | 100 |
| **Genetic/inherited disease in family** |  |  |  |
| Yes | 737 | 19.3 | 19.6 |
| No | 2182 | 57.1 | 56.8 |
| Unsure/don't know | 821 | 21.5 | 21.8 |
| Prefer not to say | 84 | 2.2 | 1.8 |
| Total | 3824 | 100 | 100 |

With regard to awareness of gene editing, most consider themselves knowledgeable (21%) or aware with limited knowledge (68%); only 6% reported having never heard of human gene editing before the survey. The level of knowledge about gene editing among the respondents is fairly high: 50 per cent of the respondents respond correctly to at least 4 of the 5 questions measuring factual knowledge about genetics and gene editing. At the conclusion of the survey, most participants reported feeling very (29%) or somewhat confident (48%) in expressing their views about gene-editing on the survey.

### Views on gene editing

Table 2 and 3 provide frequency responses to questions assessing general attitudes towards gene editing and to what degree respondents supported or disagreed with specific applications of SGE and GGE. There was a wide range of opinion on all the attitudinal questions, with many participants responding in a neutral manner. Approximately half (48%) of participants felt that if gene editing was safe it should be offered; 20% were neutral on this point. Fifty-nine percent support its availability if no other treatment options exist. Thirty two percent would consider it for their children and 29% would consider it for themselves. Respondents generally endorsed statements that each individual should have the right to make decisions about gene editing as long as the changes were not passed on (67%), and that making decisions for future generations is problematic (58%); 60% did not feel that parents had the right to edit their children’s genes before birth (T2). If gene editing is available in other countries, 35% felt it should also be available in Switzerland.

**TABLE 2.**
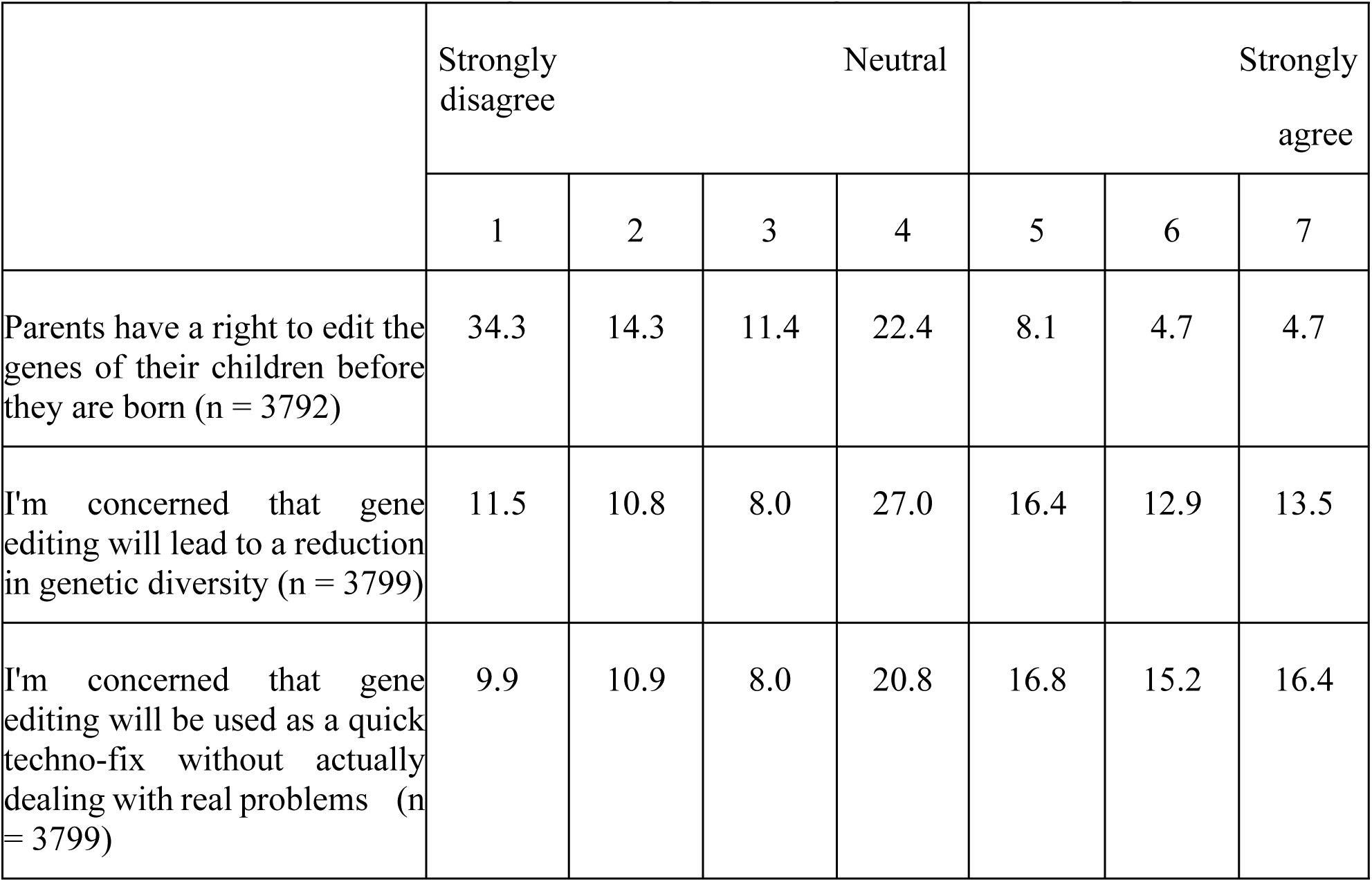
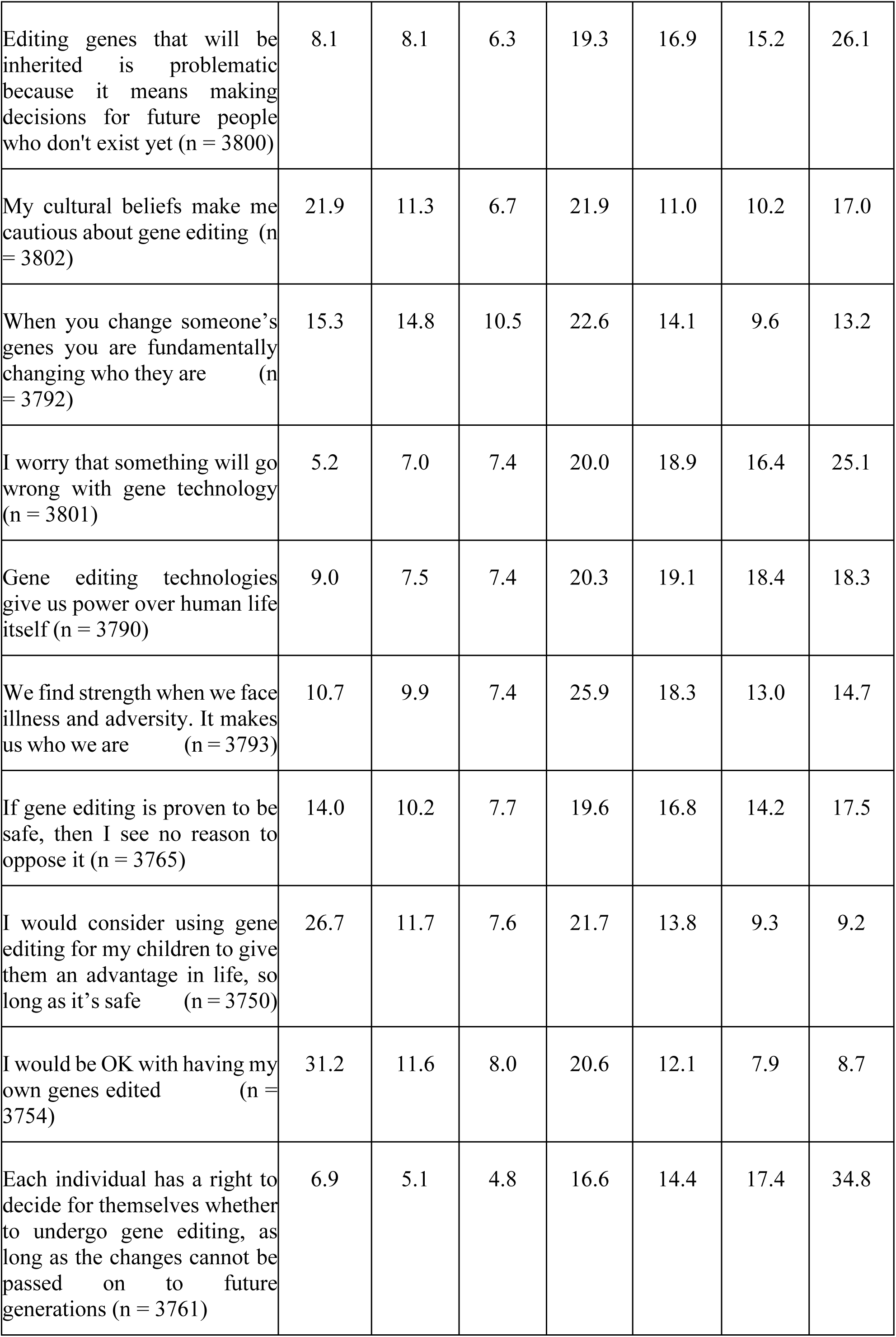
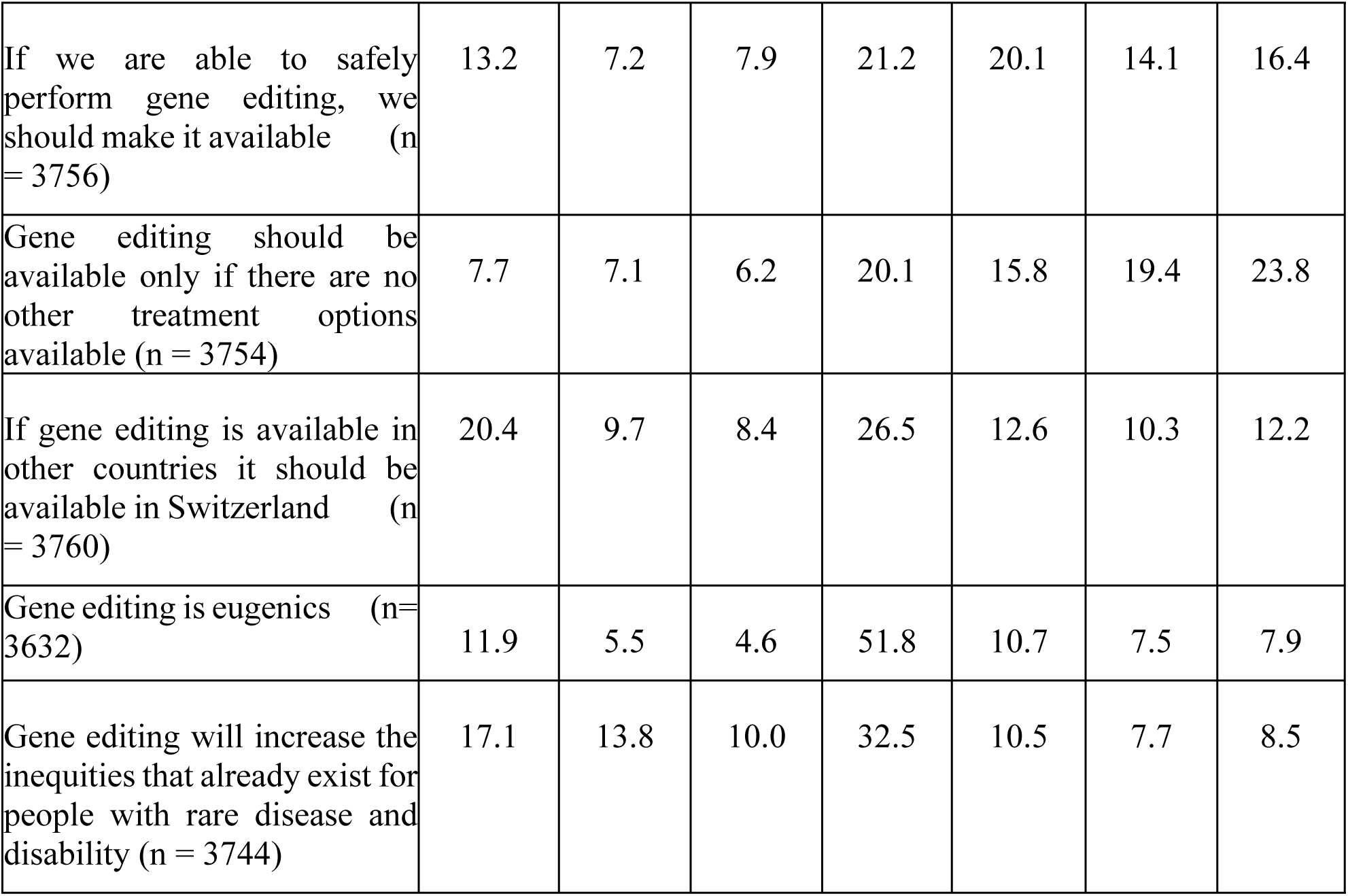
Attitudes towards gene editing (percentages in weighted sample)

When asked about which of 7 possible hypothetical uses of SGE they would support (T3), 7% disagreed with all therapeutic SGE options, 7% were neutral on all options, and 47% supported either 6 (14%) or all 7 options (33%). A majority of the Swiss public agreed with SGE for life threatening disease (76%) or debilitating disease (68%), and this agreement lessened for germline treatment of the same (57% and 45% respectively) or for more minor physical, learning or sensory impairments. Many hoped gene editing would treat or cure many conditions - the most named conditions were cancer, dementia, psychiatric illness, and conditions described as ‘severe’, ‘causing suffering’, ‘uncureable’ or those without other treatments.

*Improving human life, reducing physical and mental suffering. (original language French)*

*I think it would be good to combat life-threatening diseases such as cancer. Many people are affected by it and suffer from it and it would be revolutionary if it worked. (original language German)*

Participants were asked to what degree they endorsed potential concerns. A moderate number agreed with concerns about safety (‘something going wrong’: 60%), reduction in genetic diversity (43%), fundamentally changing who a person is (37%), or that gene editing would exacerbate already existing inequalities (27%) or was eugenic (26%). Open ended comments elaborated, including many comments that treatments are not ‘natural’.

*Human beings are imperfect and so they must remain. Unique and special. (original language Italian)*

*I think people shouldn’t interfere with nature. One day we will no longer be humans but creatures. (original language German)*

*I don’t think it’s up to our generation to decide which genes will be better for future generations. (original language French)*

Others suggested a fear of commercialization or lack of regulation:

*Do we really want to give corporations even greater control over humanity? The result*

*will be accelerating global inequalities. (original language French)*

*I see a danger in the inability to really control what goes on in laboratories in countries under authoritarian regimes where it could get out of control and generate an irretrievable drift of humanity. (original language Italian)*

Gene editing for enhancement was supported by less than 10% of respondents (T2) in either SGE or GGE. Open ended comments elaborated, frequently likening such enhancement gene editing to ‘playing god’:

*I am against playing God, not everything that is possible must be allowed. I am against parents being able to put together their children like from a catalog. (original language German)*

*Gene editing may only be used to cure or prevent serious diseases, never to change characteristics such as strength or intelligence. Keyword “superhuman”. (original language German)*

When participants were asked to endorse a list of factors that may have influenced their views towards gene editing, the most endorsed factors (nearly always citing a positive influence) included: what it means to ‘have a good life’ (57%), views towards illness and suffering (53%), views towards science and biology (50%), and what it means to be a parent and have children (49%). Most (62%) felt their personal views towards religion had a neutral influence on views towards gene editing. Open ended comments confirmed many of these factors.

*If it’s possible to modify to make life easier for someone in difficulty, or to prevent a complicated life, then I’m all for it (original language French)*

### Predictors of views on gene editing

To understand which individual factors predict the attitudes described above, we conducted multivariate analyses examining individual determinants of both broader attitudes toward gene editing and support for specific applications. Supplemental Table 2 describes the demographics of the sample used for this regression analysis (N=3429).

### Predictors of support and caution towards human gene editing

For views on gene editing under specific circumstances, the factor analysis uncovered two latent concepts in the data: one variable indicating support of gene editing, and another one expressing caution about gene editing (see Figure 1a-b).

**Figure 1a:**
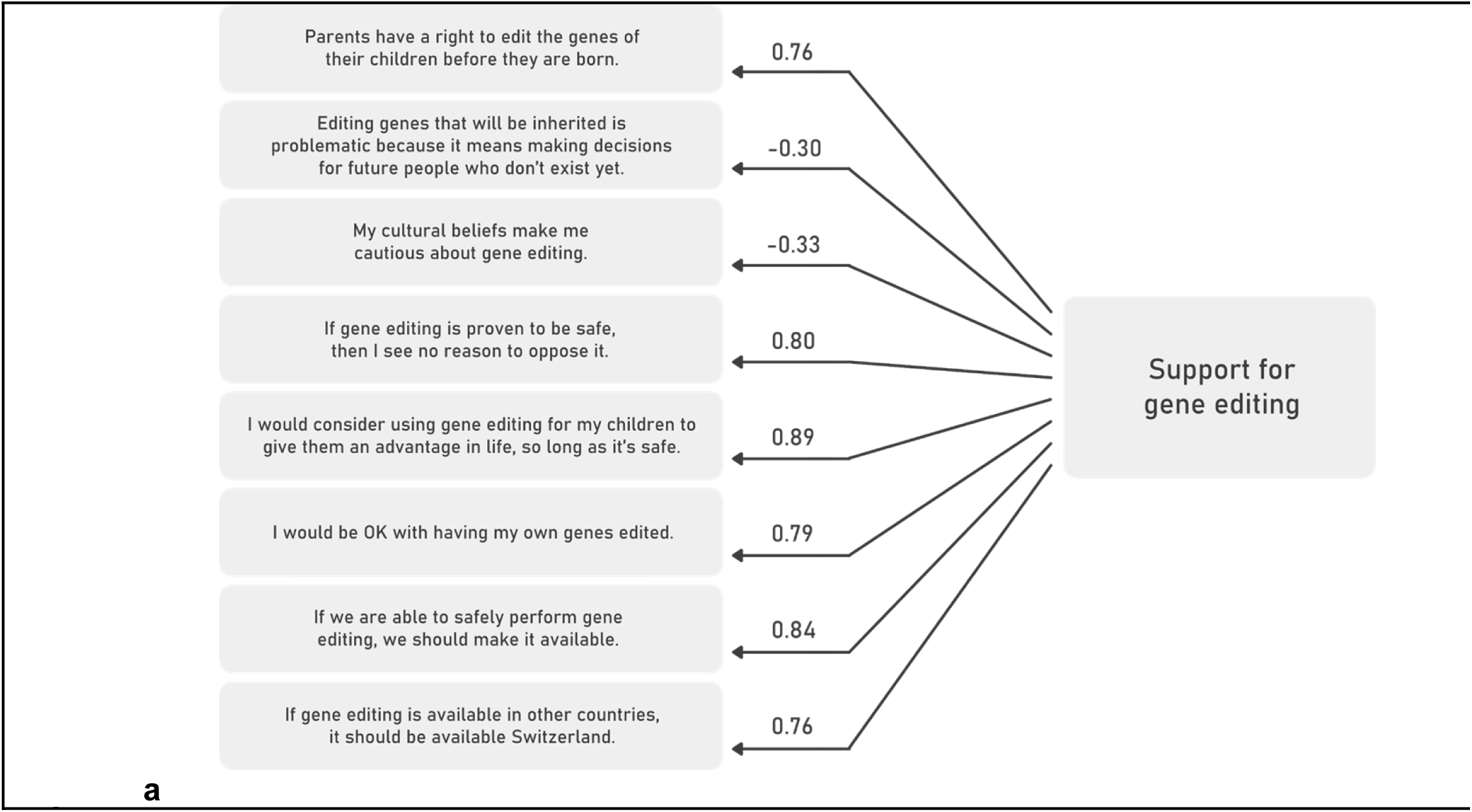
Factor structure and item loadings for support for gene editing, n=3505.

**Figure 1b:**
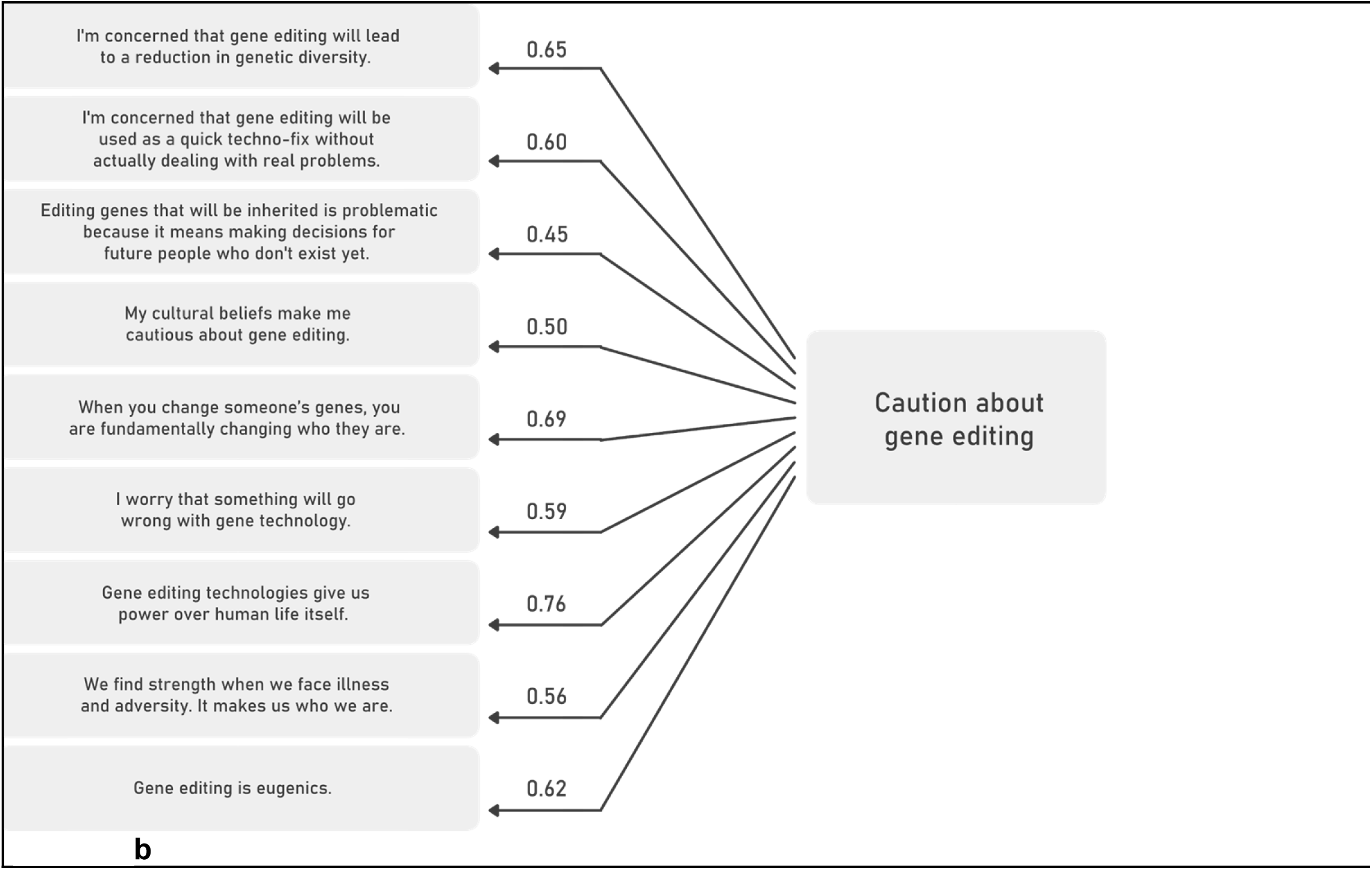
Factor structure and item loadings for caution about gene editing, n=3505.

Figure 2 presents the relationships between various individual variables and these support or caution factors (ordinary least squares regression analysis). Several demographic patterns emerge consistently across both factors. Gender is a strong predictor of views on gene editing, with female respondents showing significantly lower support and also more caution about gene editing (both p<0.001) compared to male respondents. Education also has a consistent effect, with tertiary education being associated with higher support (p<0.050) and lower caution (p<0.001) compared to secondary or other levels of education.

**Figure 2:**
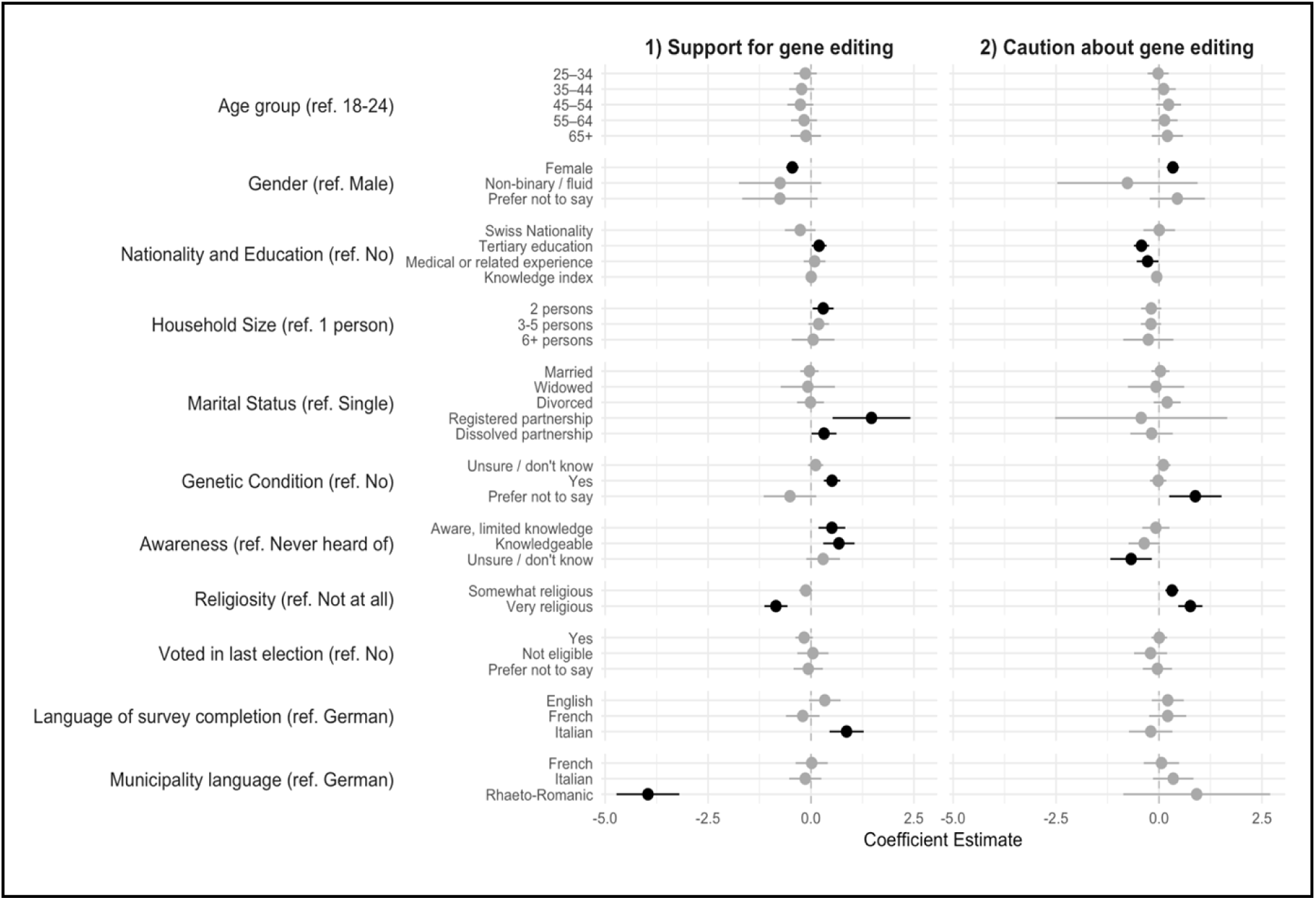
Predictors of Support and Caution Toward Gene Editing, n=3429. *Note:* Coefficients and 95% confidence intervals from weighted linear regression models. Statistically significant coefficients are bold.

Religiosity appears to have a more nuanced relationship with both attitudes. Very religious individuals showed significantly lower support for gene editing compared to those respondents who report they are not religious at all (p<0.001). For caution, we find that both moderate and strong religiosity is linked to higher levels of caution (both p<0.001), with being very religious having the strongest effect.

Personal relevance factors were especially important for support. Individuals who indicated that they or their family members have an inherited genetic condition showed significantly higher support (p<0.001) compared to those who did not report any of these conditions, as did those who self-reported awareness or knowledge of gene editing (p<0.01 and p<0.001 respectively). Experience in a medically or related field is linked to a lower level of caution (p<0.05). Regional differences were also evident, with Italian-speakers more supportive of gene editing than German-speakers (p<0.001) and respondents living in a municipality with Rhaeto-Romanic as the official language were less supportive (p<0.001).

### Predictors of views towards human gene editing uses and applications

In a next step, we examined individual predictors of support for specific uses of gene editing. Through factor analysis, we identified two measures (see figure 3a-b) for views on potential uses of gene editing: views on therapeutic gene editing and views on enhancement applications. The factor analysis revealed that respondents did not distinguish between SGE and GGE in either application. Figure 4 displays the results of the linear regression analysis. Consistent patterns across applications mirrored those for support and caution towards gene editing. Gender remains a strong predictor, with female respondents reporting a significantly lower level of agreement for both therapeutic (p<0.001) and enhancement (p<0.001) uses. Regional differences persisted, with respondents from Rhaeto-Romanic municipalities showing significantly less support for both uses of gene editing (both p<0.001) than German-speaking municipalities.

**Figure 3a:**
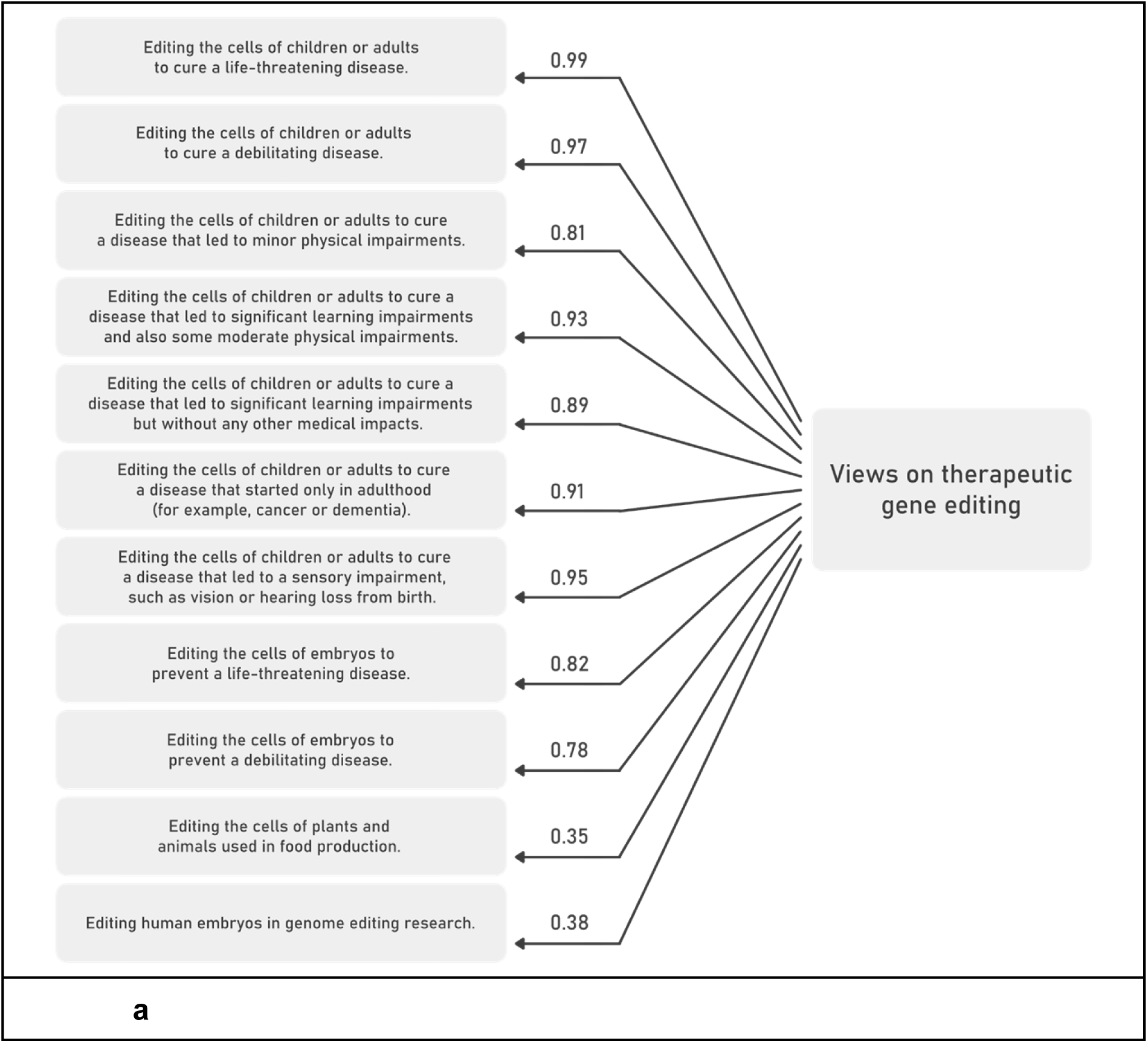
Factor structure and item loadings for views on therapeutic gene editing, n=3587.

**Figure 3b:**
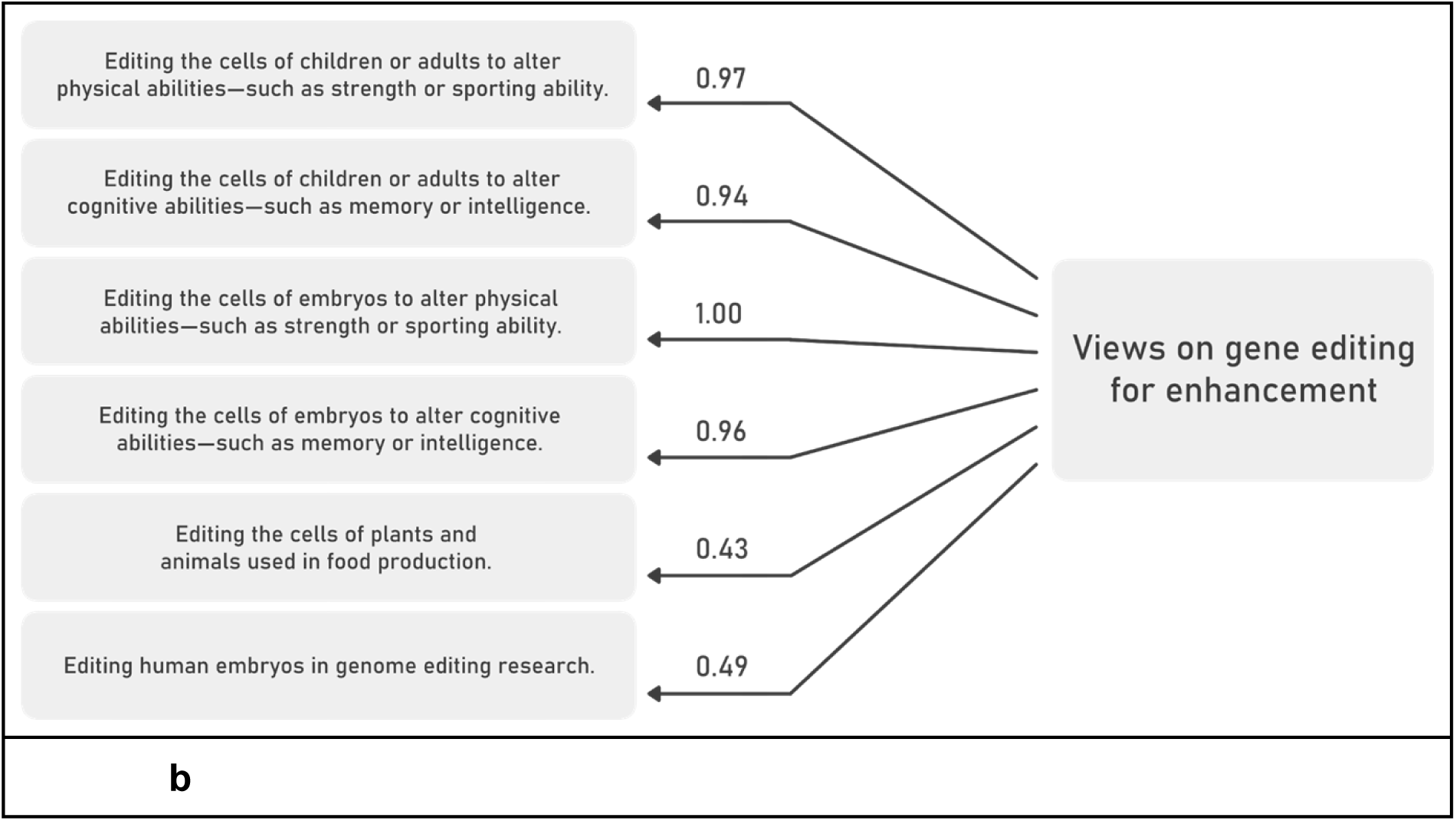
Factor structure and item loadings for views on gene editing for enhancement, n=3587.

**Figure 4:**
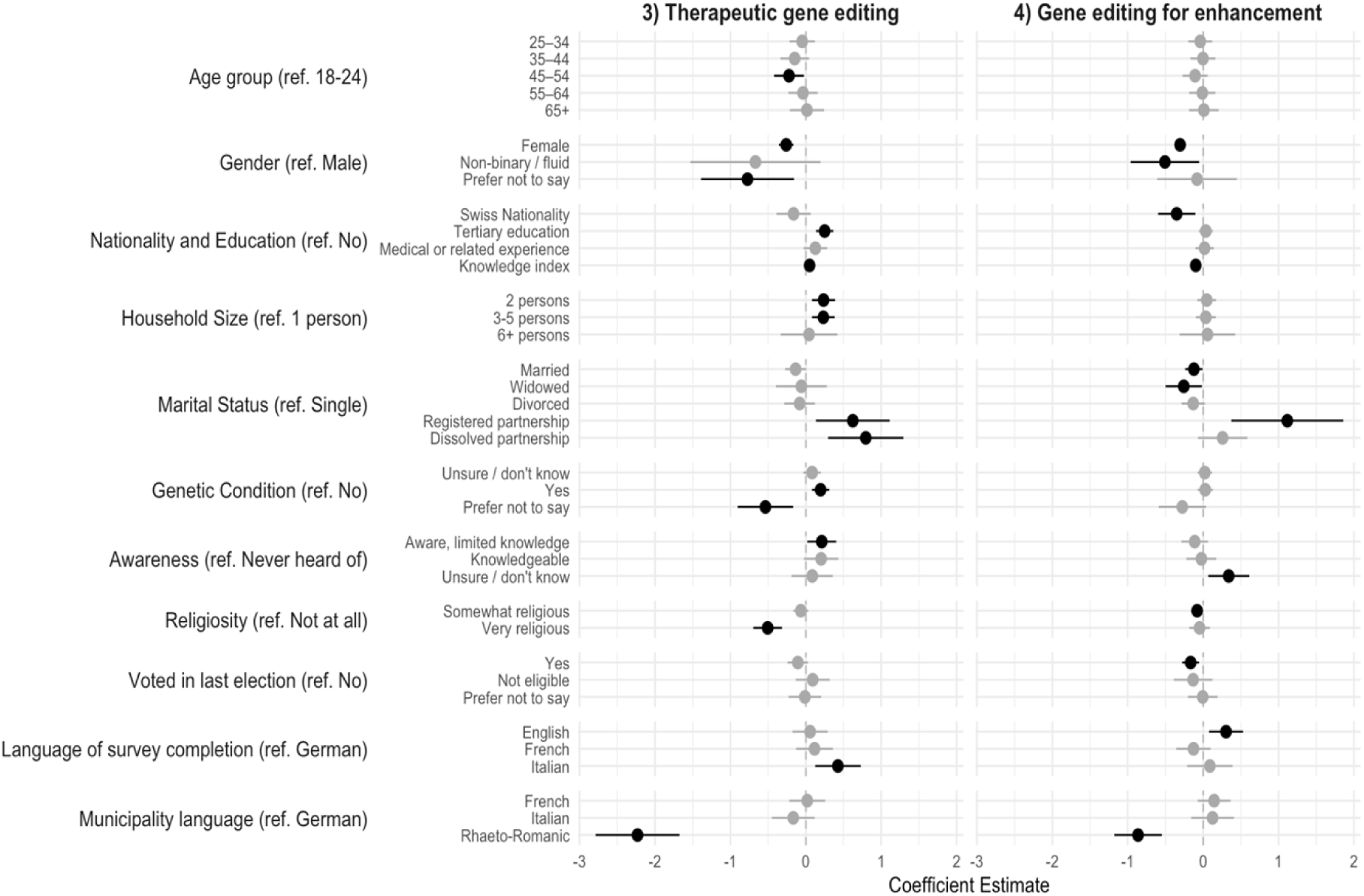
Predictors of Views on Therapeutic Gene Editing and Gene Editing for Enhancement, n=3490. *Note:* Coefficients and 95% confidence intervals from weighted linear regression models. Statistically significant coefficients in bold.

Knowledge showed contrasting effects across the two types of uses: Higher knowledge on gene editing was positively related to support for therapeutic gene editing (p<0.050) but negatively for enhancement (p<0.001), indicating that those more knowledgeable were skeptical of enhancement.

For therapeutic gene editing specifically, several additional predictors emerged. The age group between 45 and 54 was significantly less supportive than the 18-24 year olds (p<0.050). Larger household sizes (2 to 5 household members) (p<0.010), tertiary education (p<0.001), personal relevance (p<0.001) and awareness (p<0.050 ), as well as Italian-language completion of the survey (p<0.010), were all related to higher support for therapeutic gene editing. Very religious respondents showed lower support (p<0.097).

For enhancement use, the predictors differed notably. Our analyses found that marriage (p<0.050), political engagement, measured as voting in the last election (p<0.010), and Swiss Nationality were all negatively related to views on gene editing for enhancement. Non-binary/fluid gender identity, similar to being female, was related to a lower level of agreement (p<0.050). English-speaking respondents showed higher support for uses of gene editing for enhancement than German-speaking respondents (p<0.010).

### How should Swiss policies be formulated about gene editing?

We asked participants about whose voices should be heard in order to formulate policies on gene editing. Groups that were seen as completely necessary to the discussion included Swiss experts (scientists, doctors, lawyers and bioethicists; 45%), people with inherited diseases (32%) and ordinary people (30%). If a citizen jury would be held, it was most important to survey respondents that it includes presentations from experts on gene editing (47%) and from people who have lived experiences of the conditions that might potentially be treated (45%). Statements or input from international organizations (e.g. UN, WHO, OECD or EC) were seen as less necessary than hearing specifically from these Swiss groups. Finally, many of the open-ended comments at the end of the survey expressed the desire for the Swiss public to be further educated and engaged in discussions about gene editing.

*Since the political debate hasn’t yet been conducted at a societal level, the individual positions are still unclear and lacking in differentiation. (original language German)*

## Discussion

Our study documents a representative survey of Swiss residents in regards to their support and reasonings towards gene editing in Autumn 2023. This survey finds that Swiss people are moderately supportive of gene editing. We found slightly higher support of SGE than GGE, highest support for treating more serious conditions, and significantly less support for enhancement gene editing at any stage. Support was higher in those who had a family member with an inherited condition, and our data strongly supports a role of knowledge and awareness in views on gene editing for therapeutic applications. Those who were more cautious towards gene editing were more likely to be women or to have stronger self-reported religiosity. Only 7% hypothetically rejected all forms of therapeutic SGE.

When we compare the current study data with previously published papers, we note many consistencies with the existing literature. For example, the Swiss report moderate decreases in support (15-20% less) for treating similar indications when performed in embryos, although importantly both SGE and GGE views did not divide into separate factors in our analysis. This differentiation between somatic and germline is consistent with prior studies^1,7–9^. Additionally, the moderately high acceptance of SGE approaches is consistent with the expectations we had based on our prior study of Swiss professionals^23^. In the current study we noted that only 19% of participants supported gene editing research on embryos (Table 3), while approximately half voiced support for various forms of therapeutic GGE. This finding is interesting in several ways – Swiss law does not allow embryo research and historically Swiss have been slower to approve assisted reproductive technologies (for example IVF/PGD). The current data suggests that the Swiss public is more receptive to the notion of GGE than Swiss experts were^23^, but not to the degree that a referendum to change current legal restrictions would be successful in the near future. Our data did not allow us to clearly determine if the Swiss public was truly more accepting of GGE (for example because they valued stopping transmission of a genetic condition to future generations) or simply did not understand important factors about the technology. Certainly there were many open ended comments expressing that respondents felt their understanding was limited. We also note that study participants generally endorsed statements that supported the rights of children and future generations to decide for themselves about gene editing (Table 2), which is consistent with a European view towards the UN Convention on the Rights of the Child^25^, and specifically the ‘right to participate’.

**TABLE 3:**
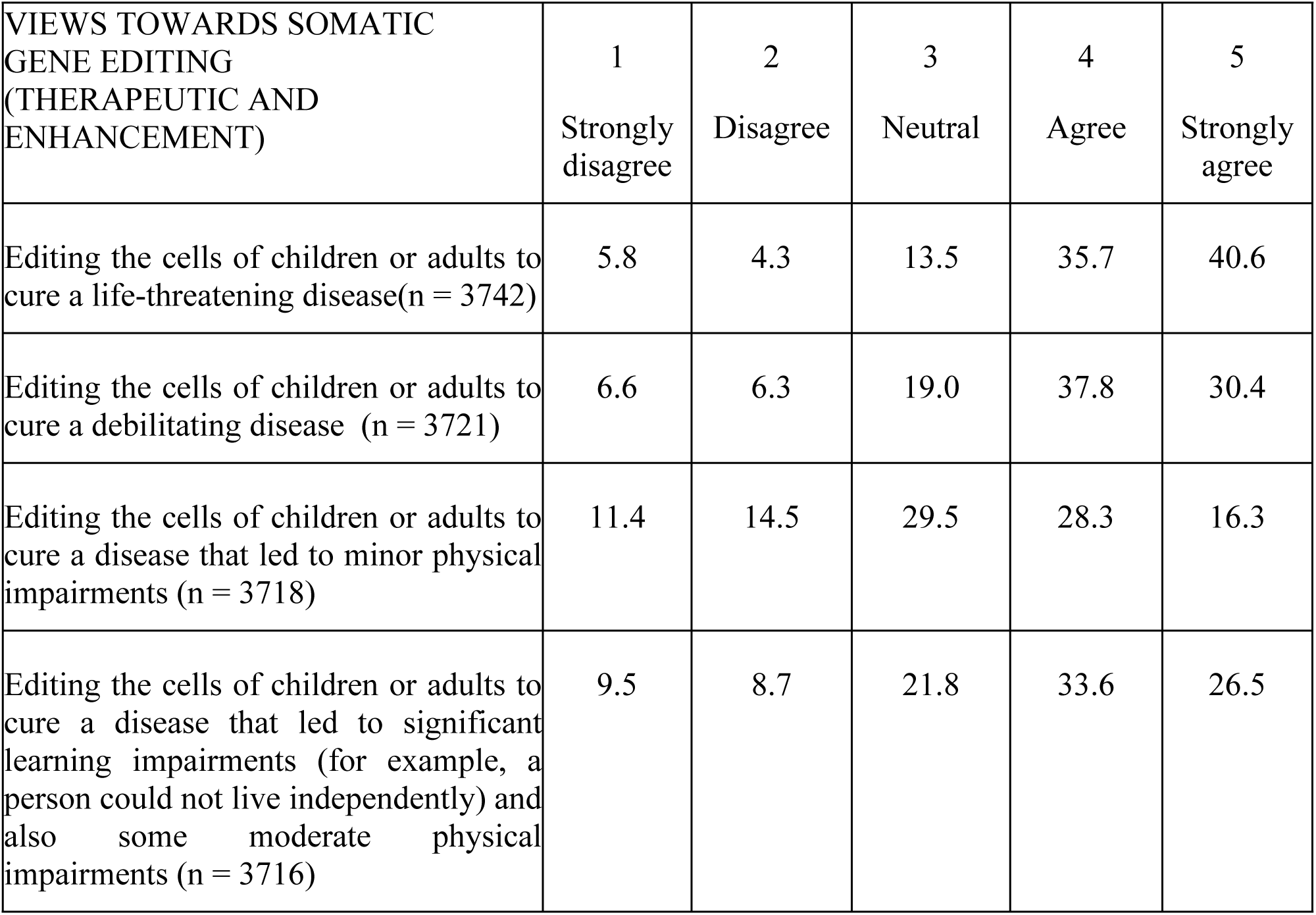
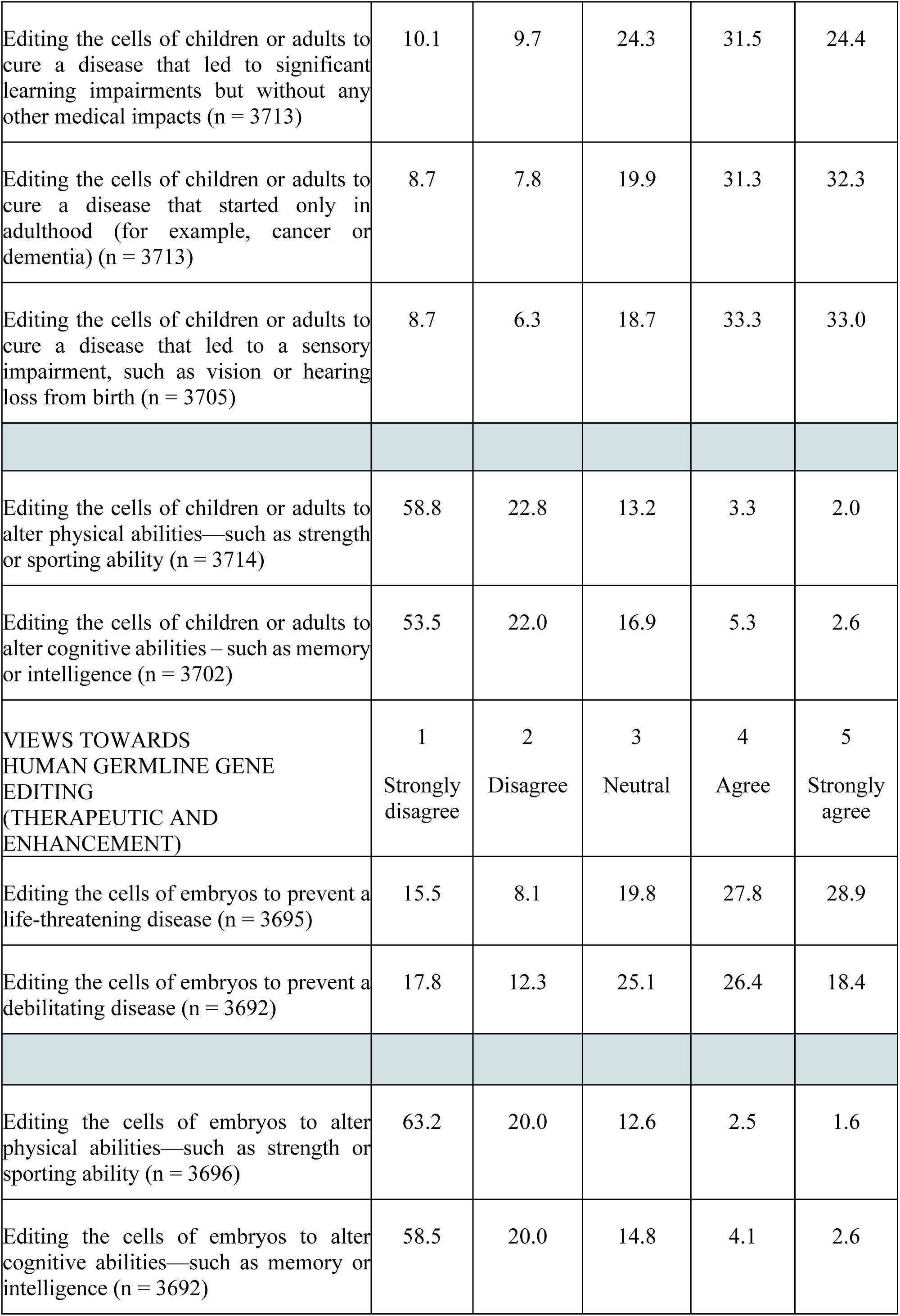
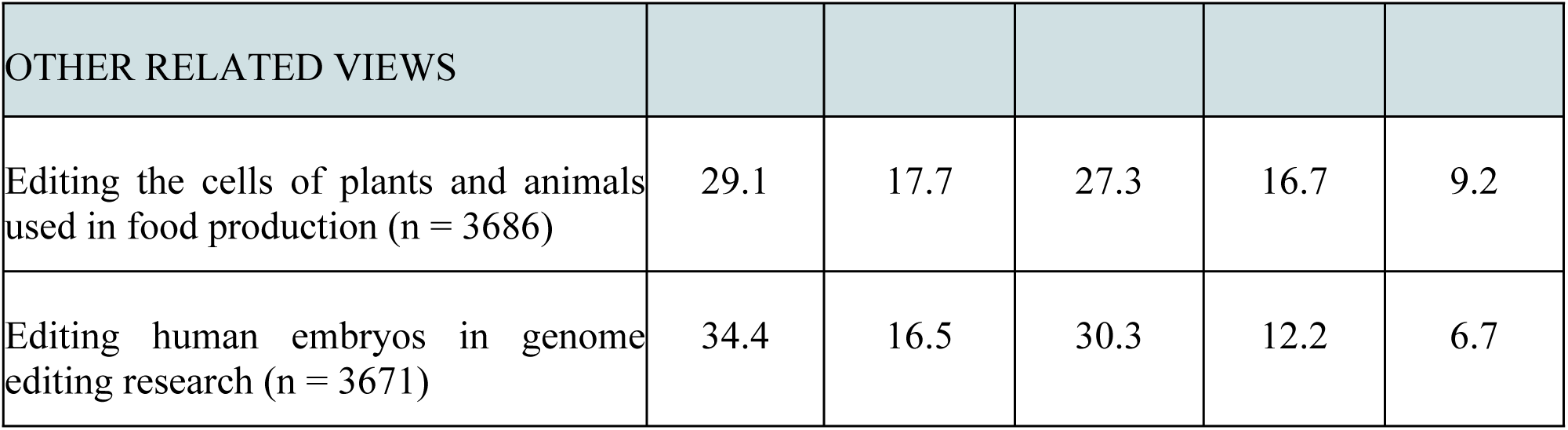
Agreement with various forms of gene editing (percentages in weighted sample)

It was not surprising to find that support for gene editing increases with disease severity and decreases when considering enhancement as these things have been consistently reported in prior literature^2,4,6,8,11–13^^4–7.^ Our study also found that respondents who indicate that they or their family members have an inherited or genetic condition express significantly higher levels of support for human gene editing than respondents reporting not to have any of these conditions. This finding is interesting as most studies of the general public suggested this did not influence views towards gene editing^1^, but studies of patient and family stakeholders identified specifically around genetic disease groups showed higher interest^4,17–21^.

Our study results were also consistent with prior literature in showing that women and self-reported very religious individuals show lower support of gene editing^1,5,6,8,10,12,13,16^, particularly in regards to GGE^3^. We note that in our prior study with Swiss experts^23^, it was predicted that religion would not be a major factor for Swiss persons in their attitudes towards gene editing, and this was also consistent, since in fact most respondents to this survey felt that religion was a neutral factor in their views. We also found that higher education (generally) and knowledge about gene editing are also positively related to higher support and lower caution towards at least therapeutic gene editing^1,6,12,16^. Finally, our study showed differences in acceptance of various forms of gene editing across the language regions of Switzerland. This reinforces our belief that even within a single country there will be local cultural factors that influence how people view gene editing.

### Limitations

Our response rate was slightly lower than 30%, although the population is generally representative of the Swiss population with the exception that it over-represents the Italian speaking region (Ticino) (33% responses, versus 4% of the Swiss population), and an under- represents the German speaking region (46% vs. 62%). However, our analysis standardized for these variations, so we expect the impact on our results and interpretation is minimal. It is certainly possible that those with stronger opinions (either for or against) were more likely to respond to the survey, however.

### Conclusions

This study documents the views of the Swiss public towards SGE and GGE. It reveals moderate support for therapeutic SGE, with decreasing support as the conditions become less serious or move towards enhancement. Most reasons for hypothetically supporting gene editing treatments relate to views towards what it means to live a good life, and views towards science and medicine, rather than religious views. Our study findings map to those documented in other countries in some, but not all cases.

## Data Availability

The survey data (minus the confidential data from the FSO) will be available upon reasonable request from the authors. Qualitative data (unlinked from survey data) will be available upon reasonable request from the authors. The code will be available upon reasonable request from the authors.

## Data Availability Statement

The survey data (minus the confidential data from the FSO) will be available upon reasonable request from the authors. Qualitative data (unlinked from survey data) will be available upon reasonable request from the authors.

## Code Availability

The code will be available upon reasonable request from the authors.

## Financial Disclosure Statement

The author(s) received no specific funding for this work.

## Acknowledgements

We appreciate the translation assistance and skills of the following colleagues and interns in the Health Ethics and Policy Lab: Agata Ferretti and Mattia Andreoletti (Italian), Jade Berlincourt, Marc Reynaud and Eirini Petrou (French), Constantin Landers, Sara Kijewski, Naomi Wyler, Joanna Sleigh, Martina Bredemeyer and Corinne Strässle (German). Thanks to Stefan Wehrli, Patricia Wäger, Paola Gagliardi and David Presberger at the ETH Decision Science Laboratory for their significant assistance in administering the survey and additional translation assistance. Thanks also to Han (Ray) Yi for help with early visualizations and assistance in cleaning the data. Significant thanks to Simon Niemeyer, Dianne Nichol and their colleagues in the Australian Citizens Jury, who shared their survey measure for adaption in this project.

## Author Contribution Statement

● Conception or design of the work: KO, EV
● Development of the measure: KO, SK, EP, AF. Translation assistance as listed in acknowledgements.
● Acquisition of data: KO, EP
● Analysis, or interpretation of data for the work: KO, SK, NW, AF, EP, CR
● Drafting the work: KO, SK, NW
● Visualizations: NW, SK and HY.
● Supervision of project: KO and EV.
● Funding: EV (lab funding).
● Reviewing it critically for important intellectual content and final approval of the versions to be published: KO, SK, NW, AF, EP, CR, EV
● Agreement to be accountable for all aspects of the work in ensuring that questions related to the accuracy or integrity of any part of the work are appropriately investigated and resolved: KO, SK, NW, AF, EP, CR, EV

## Funding

No external funding was received for this project.

## Ethical Approval

The survey has been approved by the ethics commission of the ETH Zurich (EK 2022-N-84). Participants provided their informed consent at the start of the online survey.

## Competing Interests

The authors declare no competing interests.

## Summary of supplemental files

Supplemental Methods: Includes operationalization of variables (Supplemental Table S1) and English survey measure.

Supplementary Table 2: Participant demographics - regression analysis sample.

Supplementary Table 3: Predictors of views on human gene editing (OLS Regression results).

Supplementary Table 4: Predictors of views on potential uses of gene editing (OLS regression estimates)

Extra copies of Figures (in case they do not come through properly in the main text)

## Notes

### Competing Interest Statement

The authors have declared no competing interest.

